# Obesity is associated with greater variability of reward signals in the nucleus accumbens

**DOI:** 10.1101/2025.09.04.25335092

**Authors:** Mechteld M. van den Hoek Ostende, Anne Kühnel, Monja P. Neuser, Thomas Dresler, Jennifer Svaldi, Nils B. Kroemer

**Author notes:** **Corresponding author:** Prof. Dr. Nils B. Kroemer, Venusberg Campus 1, 53127 Bonn, Germany.

## Abstract

**Background:** Binge eating disorder (BED) is characterized by repeated episodes of binge eating accompanied by a loss of control. Although it remains unknown which factors drive binge eating (BE) episodes, there are indications that variability in nucleus accumbens (NAcc) responses could lead to increased variability in food intake.

**Methods:** Here, we assessed whether BED is associated with higher intra-individual variability in behavioral and neuroimaging indices of reward responses. To this end, patients with BED (*n* = 35, *M*_BMI_ = 33.2 kg/m^2^ ± 6.8), participants with subsyndromal BED (*n* = 21, *M*_BMI_ = 29.0 kg/m^2^ ± 7.2), and individuals without symptoms of binge eating (*n* = 23, *M*_BMI_ = 32.3 kg/m^2^ ± 6.5) completed an effort allocation task with concurrent functional magnetic resonance imaging.

**Results:** In line with our hypothesis, we found that patients with BED had higher variability in subjective wanting ratings of food (*F*_34,21_ =1.48, *p*_boot_ = .024), but not effort exertion (*F_34_*_,21_ = 1.13, *p*_boot_ = .30). Crucially, trial-by-trial variability in NAcc responses during the presentation of cues was associated with a higher BMI (*b*=0.11, 95%CI [0.03, 0.19], BF_10_=11.1) and disinhibited eating (*b*=0.19, 95%CI [0.01, 0.36], BF_10_=4.0) across groups, whereas NAcc variability was only marginally elevated in patients with BED (*b*=0.12, 95%CI [−0.04, 0.29], BF_10_=1.2, P>0|data = 88%).

**Conclusions:** Our results support the idea that BMI and disinhibited eating are associated with more variable NAcc responses, which may contribute to the symptoms of BED. However, this association is only weakly indicative of clinical severity of BED.

## Introduction

Binge eating disorder (BED) is the most prevalent eating disorder (1), affecting approximately 1.5% of women and 0.3% of men (2). BED is characterized by repeated binge eating (BE) episodes, in which an objectively large amount of food is consumed, accompanied by experienced loss of control. Individuals with BED present with high rates of overweight and obesity (3), and mental and physical comorbidities (2, 3). Whereas circumstances under which BE are more likely to occur have been documented (e.g., 4, 5, 6), the mechanisms underlying BE remain largely elusive.

Cognitive and motivational models of eating behavior posit that attentional biases are important in maintaining eating disorders including BED (e.g., 7, 8-10). Such biases may be rooted in incentive sensitization (11); through frequent encounters with food rewards, the motivational pathway is sensitized to food cues. Consequently, future encounters are associated with higher arousal and craving (9, 12). In line with incentive-sensitization theory, fMRI studies found increased activation of the mesocorticolimbic system to food in individuals with BED (13, 14) while activity in regions that control food intake, such as the dorsolateral prefrontal cortex (dlPFC), is reduced (15). Moreover, simulations of food-related reward behavior demonstrate that fluctuations in food reward responses could explain an increased variability in food intake (16). Thus, variability in reward sensitivity could provide a more nuanced model of the contribution of reward processing to the etiology of BED.

To better conceptualize the inherent variability of eating behavior, intra-individual variance—in addition to differences in response amplitude—of behavioral and neural indices of reward processes is highly promising (16). Since phases of food intake alternate with phases of fasting (17, 18), BE episodes or excessive restraint may be reconceptualized as extremes of a distribution. In other words, if homeostatic signals that regulate food intake are overridden through hedonic eating (“disinhibited”), this variance increases. Thus, episodes of disinhibited eating and BE may conceivably increase variability (16). This idea is supported by an association of the variability in the nucleus accumbens (NAcc) to milkshake receipt with greater variability in subsequent *ad libitum* food intake in a sample of participants without pathological eating behavior (19). In this study, variable NAcc responses were associated with increased body mass index (BMI) and disinhibited eating. Hence, higher intra-individual variability in reward sensitivity may be associated with higher variability in food intake, including variability caused by BE episodes.

Conceptually, the effects of elevated variability in reward sensitivity should extend beyond food intake. Viewed as an economic exchange, willingness to work for a specific reward reflects the reward’s value (20, 21); as the reward value increases, the effort to obtain it increases. In other words, the motivational processes that drive behavior to obtain (food) rewards are mediated by reward sensitivity through cost-benefit computations (21, 22). Effort allocation tasks (EAT) capture such tradeoffs since the decision to exert effort is influenced by the dynamic integration of its perceived benefits against its perceived costs (i.e., physical force; 22, 23-25). Since the expected reward value underlying these computations is not a set value, but rather a distribution of possible outcomes (26), fluctuations in the reward value account for a substantial variance in subsequent decision-making (27–29). In line with this prediction, BMI was associated with higher variability in subjective wanting ratings (30). Accordingly, animal studies demonstrate that uncertainty of reward receipt could exacerbate such variability; when rodents are provided food at insecure feeding schedules, binge-like eating behavior ensues (31, 32). This uncertainty in availability was also associated with weight cycling (33). As such, variability in the motivation to work for reward should be most pronounced when uncertainty about the difficulty—and therefore receipt of reward—is introduced in the parameters of the cost-benefit computation.

Based on previous evidence showing that altered reward processing is implicated in the etiology and maintenance of BED, we determined whether BED is associated with increased variability in reward sensitivity using comprehensive behavioral and neural indices of reward sensitivity. We indexed the variability of the reward value of food and monetary rewards in individuals with BED, subsyndromal BED and weight-matched controls. We predicted that variability in task behavior reflects variability in neural reward signaling (19). We expected variability to be exacerbated if the effort required was uncertain and manipulated the uncertainty of the effort requirements accordingly. We hypothesized that individuals with BED are characterized by increased variance in reward sensitivity, which is reflected in increased variability in effort exertion and in NAcc responses. Moreover, we expected increased BE symptomology to be associated with increased variability in reward sensitivity.

## Methods

### Participants

In total, 79 women participated in the first session, of which 35 fulfilled the criteria of BED (range BMI: 22.6 – 54.6), 21 experienced subsyndromal binge eating (subBED, range BMI: 20.1 – 42.5), and 23 individuals did not experience binge eating (no BE, range BMI: 22.2 – 44.9; Table 1). From this original sample, 59 participants (Figure 1D) also completed the fMRI session (several BED patients had MR contraindications and therefore only completed a behavioral session). For both sessions, groups were comparable regarding BMI and age.

**Figure 1:**
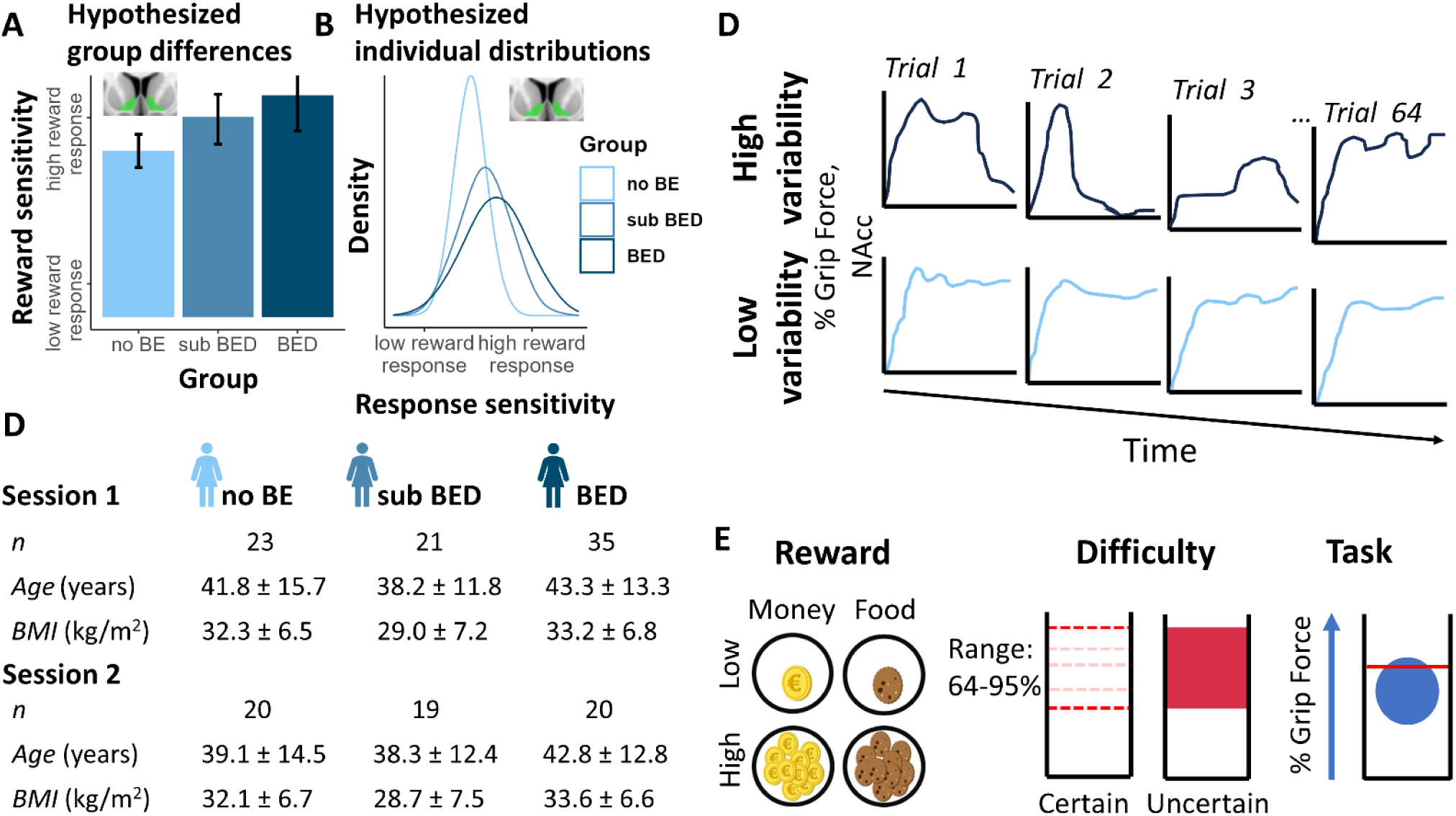
Study rationale and procedures. A: Hypothesized differences in average reward sensitivity across groups. B: Hypothesized differences in the distribution of reward sensitivity across groups. C: Schematic depiction of high and low variability over trials. D: Sample characteristics. E: Schematic overview of the grip force effort allocation task. Trials consisted of a jittered fixation cross, a preparation phase, an effort phase in which participants used grip force to move a ball in a tube, and a feedback phase (Supplemental Information). BE = binge eating, BED = binge eating disorder, BMI = body mass index, NAcc = nucleus accumbens

**Table 1.**
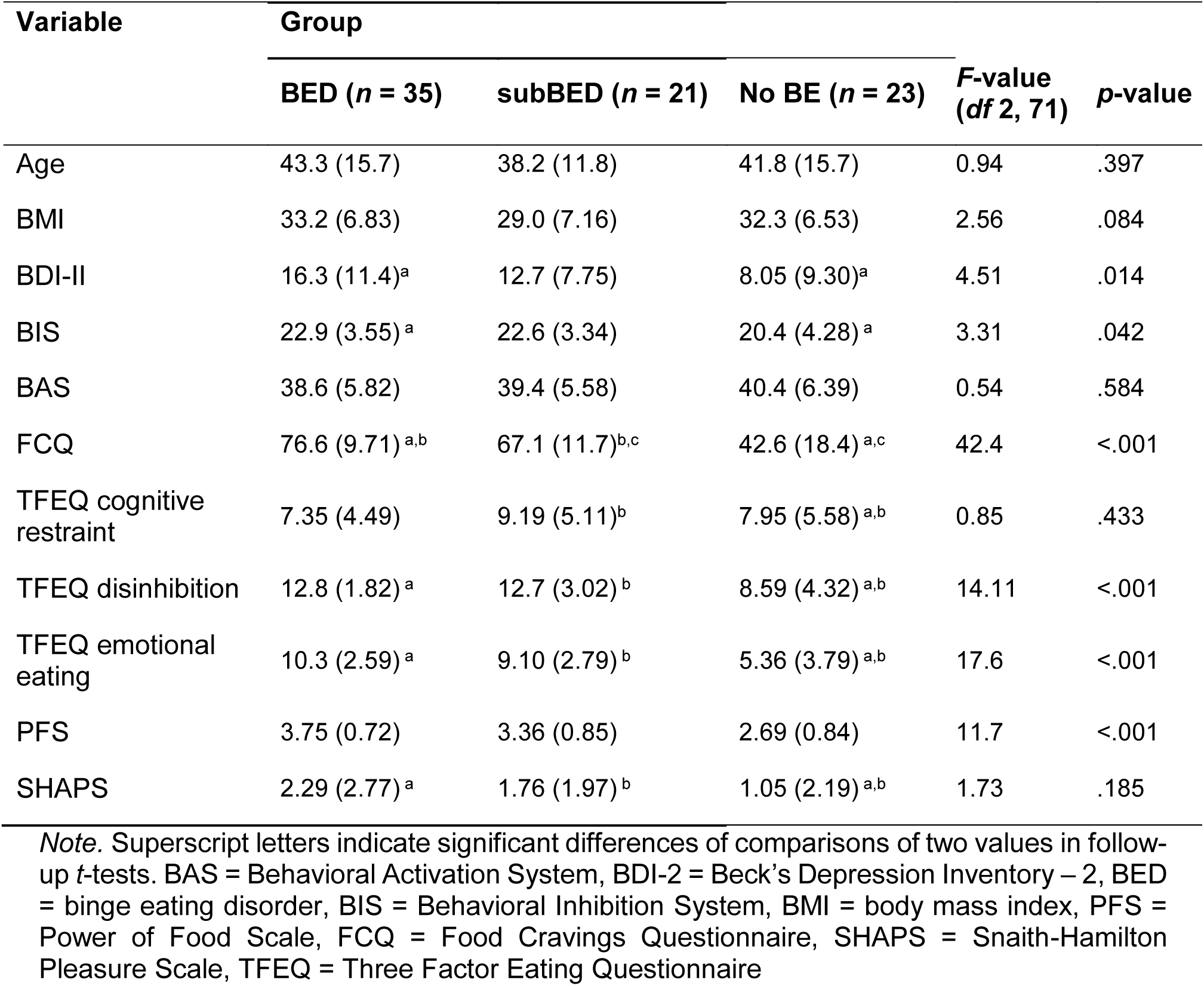
Demographic and clinical characteristics of the full sample.

| Variable | Group |  |  | *F*-value<br>(*df* 2, 71) | *p*-value |
| --- | --- | --- | --- | --- | --- |
|  | BED (*n* = 35) | subBED (*n* = 21) | No BE (*n* = 23) |  |  |
| Age | 43.3 (15.7) | 38.2 (11.8) | 41.8 (15.7) | 0.94 | .397 |
| BMI | 33.2 (6.83) | 29.0 (7.16) | 32.3 (6.53) | 2.56 | .084 |
| BDI-II | 16.3 (11.4)<sup>a</sup> | 12.7 (7.75) | 8.05 (9.30)<sup>a</sup> | 4.51 | .014 |
| BIS | 22.9 (3.55)<sup>a</sup> | 22.6 (3.34) | 20.4 (4.28)<sup>a</sup> | 3.31 | .042 |
| BAS | 38.6 (5.82) | 39.4 (5.58) | 40.4 (6.39) | 0.54 | .584 |
| FCQ | 76.6 (9.71)<sup>a,b</sup> | 67.1 (11.7)<sup>b,c</sup> | 42.6 (18.4)<sup>a,c</sup> | 42.4 | <.001 |
| TFEQ cognitive restraint | 7.35 (4.49) | 9.19 (5.11)<sup>b</sup> | 7.95 (5.58)<sup>a,b</sup> | 0.85 | .433 |
| TFEQ disinhibition | 12.8 (1.82)<sup>a</sup> | 12.7 (3.02)<sup>b</sup> | 8.59 (4.32)<sup>a,b</sup> | 14.11 | <.001 |
| TFEQ emotional eating | 10.3 (2.59)<sup>a</sup> | 9.10 (2.79)<sup>b</sup> | 5.36 (3.79)<sup>a,b</sup> | 17.6 | <.001 |
| PFS | 3.75 (0.72) | 3.36 (0.85) | 2.69 (0.84) | 11.7 | <.001 |
| SHAPS | 2.29 (2.77)<sup>a</sup> | 1.76 (1.97)<sup>b</sup> | 1.05 (2.19)<sup>a,b</sup> | 1.73 | .185 |

*Note.* Superscript letters indicate significant differences of comparisons of two values in follow-up *t*-tests. BAS = Behavioral Activation System, BDI-2 = Beck's Depression Inventory – 2, BED = binge eating disorder, BIS = Behavioral Inhibition System, BMI = body mass index, PFS = Power of Food Scale, FCQ = Food Cravings Questionnaire, SHAPS = Snaith-Hamilton Pleasure Scale, TFEQ = Three Factor Eating Questionnaire

Group membership was determined through the Eating Disorder Examination (EDE; 34). For the subBED group, all diagnostic criteria of BED according to DSM-5 needed to be fulfilled, with the exception that the frequency of BE episodes was lower than once a week for 3 months. Participants were excluded if they fulfilled the criteria for bipolar disorder, or alcohol/substance use disorder within the past six months as determined by the Structured Clinical Interview for DSM-IV (SCID; 35). Given high comorbidity rates with major depressive disorder (1), we only excluded participants with comorbid depression if they reported acute suicidality, or if antidepressive medication was started or altered in the past two months. We additionally excluded no BE and subBED participants who fulfilled the lifetime criteria for any lifetime eating disorder.

The study was preregistered (NCT04184856) and approved by the institutional review board of the University of Tübingen (3939/2017BO2) and completed in line with the Declaration of Helsinki. Participants provided written informed consent before the diagnostic interview. For the full study, participants received €110 with potential additional winnings based on task performance.

### Experimental procedure

We assessed in- and exclusion criteria through a telephone screening (∼25 min). Suitable participants subsequently partook in an online study and two laboratory sessions. The online part included questionnaires on aberrant eating behavior and psychopathology, among them the three factor eating questionnaire (TFEQ; 36), which measures cognitive restraint, dietary disinhibition and non-homeostatic eating. For the first session, we asked participants to eat approximately 1.5 h in advance (neither hungry nor full). After signing informed consent, participants completed a practice round of the grip force EAT (grEAT). Subsequently, we conducted a clinical interview. Participants then completed a food cue reactivity task (FCR, ca. 20 min; 37), before completing the grEAT (ca. 40 min; 38). Finally, participants took part in a taste test (ca. 20 min; 39). The first session lasted approximately 3.5 h.

For the second session, participants came to the lab after an overnight fast (i.e., no caloric intake >8 h prior to the session). Participants received a small breakfast and completed a reinforcement learning task (40). They completed a second grEAT (ca. 30 min) and a food bidding task (ca. 20 min) with concurrent fMRI. In total, the second session lasted approximately 4.75 h. The current paper focuses on the grEAT, while the other tasks are reported elsewhere.

### Grip force effort allocation task

In line with previous work (30), we estimated variability in reward value through its influence on effort exertion. To this end, we combined the task from Neuser, Teckentrup (25) with grip force as input modality (23) and uncertainty difficulty in a subset of trials. For each participant, we determined the maximum grip force (25) and an exchange rate for points to money/kcal was adapted according to the ratio of the linear exertion slopes of exerted force for standardized amounts of money and snacks.

In the grEAT (Figure 1E; details in the SI), participants worked for monetary and food rewards in low (one point per second) and high magnitude trials (10 points/s). To incorporate previous insights about uncertainty and BE, the difficulty was uncertain (no line shown) in half of the trials. In trials of the grEAT in the first session, participants additionally indicated on 0-100 visual analog scales (VAS) how much they exerted themselves, how much they wanted the reward.

### MRI data acquisition and preprocessing

fMRI data (∼35 min, 1,500 volumes, see SI) were acquired on a Siemens 3T Prisma scanner with a 64-channel head coil. Data was then preprocessed using fMRIprep (41) and smoothed (6×6×6 mm³). For confound correction in first-levels, we extracted the average white matter and CSF signal as well as the six motion regressors.

### Data analysis

#### Behavioral data

Using behavioral data from the first session, we assessed effort and wanting ratings. We defined effort as the percentage of the force of each participant’s individual maximum grip force. We computed the average effort per trial and used linear mixed-effect models as implemented in R (lmerTest) using group (no BE, subBED, BED), reward type (food, monetary), and reward magnitude (low, high), and their interactions as fixed effects with BMI and age as covariates. As random effects of participants, we used random intercepts and slopes for reward type and reward magnitude. To evaluate variability in two states where the costs of effort are known versus unknown, we estimated separate models for certain and uncertain trials.

In line with previous work (19, 30), we defined variability as trial-wise residuals (i.e., after accounting for systematic effects of conditions) of the relative effort exerted (“objective” value) and wanting ratings (subjective value) after each trial. We anticipated that differences in variability would show primarily in uncertain trials as they reflect the internal valuation of the rewards independent of the difficulty. Therefore, to test whether variability differed between groups, we bootstrapped (1,000 resamples) variance ratio tests (Var(BED/subBED)/Var(no BE)) on residuals of uncertain trials for food and monetary rewards (28). To assess associations with BMI, we calculated the individual SD of the residuals and report bootstrapped Pearson correlation with BMI.

#### fMRI data

Statistical analyses of fMRI data were conducted using SPM12. The first-level general linear models included regressors for cue, work, and feedback phases (see SI). For the variability analyses, we extracted the adjusted signal (filtered and corrected for covariates) in the NAcc, dlPFC, and temporal lobe control regions (anterior: aMTL, posterior: pMTL, temporo-occiptal: toMTL). Regions of interest (ROI) were defined from the Harvard Oxford atlas (42) and combined across hemispheres. The dlPFC was derived from Neurosynth (meta-analysis with the term dlPFC, *z*-threshold = 5.2, smoothed with a 4×4×4 mm³ kernel).

To calculate individual variability in anticipatory cue responses, we used linear mixed-effects models (*fitlme*, MATLAB v2023a) estimating brain responses in each trial as previously established (19). To estimate trial-wise responses, we assigned all volumes a corresponding trial number with a new trial starting once a new cue was shown. The models predicting activation in the ROIs included all task regressors from the first-level model (i.e., after convolution with the hemodynamic response function and filtering). To account for differences between trials, we included interactions of cue presentation with reward magnitude (centered), difficulty (centered), uncertainty (dummy-coded), and reward type (dummy-coded). The models included random intercepts and slopes for all predictors and a nested trial term (1 + Cue|ID:Trial) so that distinct deviations in activation from the group means are estimated for each trial and each individual.

To determine the effects of group and BMI on the variability in cue responses, we estimated Bayesian mixed location-scale models using brms with the trial residuals (i.e., deviation from the condition mean) as outcome. The model for the amplitude (location) only included a random intercept. The model for individual variance terms included the task conditions reward type, reward magnitude, their interaction and random intercepts and slopes for task conditions. We included BMI (*z*-standardized) and Group (dummy-coded) and their interaction with the task conditions as fixed effects. We estimated separate models for all 5 ROIs (NAcc, dlPFC, aMTL, pMTL, toMTL). To explore whether effects are driven by certain or uncertain trials, we estimated separate models post hoc. Bayesian models are evaluated using the 95% credible interval that does not include 0 if an effect is significant. In addition, Bayes Factors (BF) quantify the evidence for or against the null hypothesis, where BF_10_ quantifies the evidence for the (undirected) alternative hypothesis.

In the current manuscript, we are focusing on differences in the variability of the reward response, as differences in amplitude of the anticipatory NAcc response have been reported elsewhere, showing a lower reward sensitivity (i.e., difference in NAcc response to high vs. low rewards) in BED compared to no BE but no association with BMI (38). Crucially, we used the hierarchical estimation of trial coefficients chosen here and then analyzed them using the same linear mixed-effects model as for the behavioral session.

#### Statistical threshold and software

We used a two-tailed threshold of α < .05 to interpret the significance of our findings. We preprocessed the raw behavioral data in MATLAB v2021a. Linear mixed-effects models (43) were analyzed with lmerTest (44) and location-scale models to investigate changes in variability with *brms* (45) in R (46). We used *ggplot2* (47) and *ggdist* (48) for data visualization.

## Results

### No differences between groups for average effort and wanting in the session one

To characterize average effort, we used separate mixed-effects models for certain and uncertain trials of the first session. As anticipated, participants exerted more effort in trials with a large vs. small reward at stake (certain: *b* = 14.3, *t*_95.2_ = 3.09, *p* < .001; uncertain: *b* = 18.7, *t*_93.2_ = 3.33, *p* < .001). Furthermore, they exerted more effort for money vs. food rewards (certain: *b* = 12.2, *t*91.4 = 5.01, *p* = .003; uncertain: *b* = 13.9, *t*_89.0_ = 3.09, *p* = .001). Likewise, participants reported wanting large rewards more than small rewards (certain: *b* = 25.3, *t*_89.3_ = 5.48, *p* < .001; uncertain: *b* = 26.5, *t*_91.3_ = 6.00, *p* < .001) and money more than food rewards (certain: *b* = 16.0, *t*_77.3_ = 3.25, *p* = .002; uncertain: *b* = 16.0, *t*_85.9_ = 2.98, *p* = .004). No other predictors were significant (see SI; 38).

### Patients with BED have increased variability in wanting ratings of food

To evaluate differences in variability of behavior, we compared the trial-to-trial variability of effort maintenance and wanting in patients with BED using bootstrapped variance ratio tests (*F*-test). As hypothesized, in uncertain trials, patients with BED showed higher variability in wanting (Fig. 2) for food (*F*_34,22_=1.48, *p*_boot_ = .024), but not money (*F*_34,22_= 0.97, *p*_boot_ = .56). No differences were observed for the subBED group (food: *F*_21,22_= 0.95, *p*_boot_ = .62, money: *F*_21,22_= 1.00, *p*_boot_ = .52). In contrast to the self-reported wanting, effort maintenance did not vary differentially between groups (*p*s > .11, Table S1). In certain trials, there were no associations with wanting. However, individuals with subBED showed consistently higher variability in effort maintenance (*p*s < .001, Table S1) and patients with BED showed higher variability in money trials (*F*_34,22_=1.67, *p*_boot_ = .007). Against our expectation, BMI was not correlated with individual SD of effort maintenance (*r*= −.06, 95CI_boot_ [−.22, .08]) or wanting ratings (*r*= .07, 95CI_boot_ [−.08, .23], Table S2).

**Figure 2:**
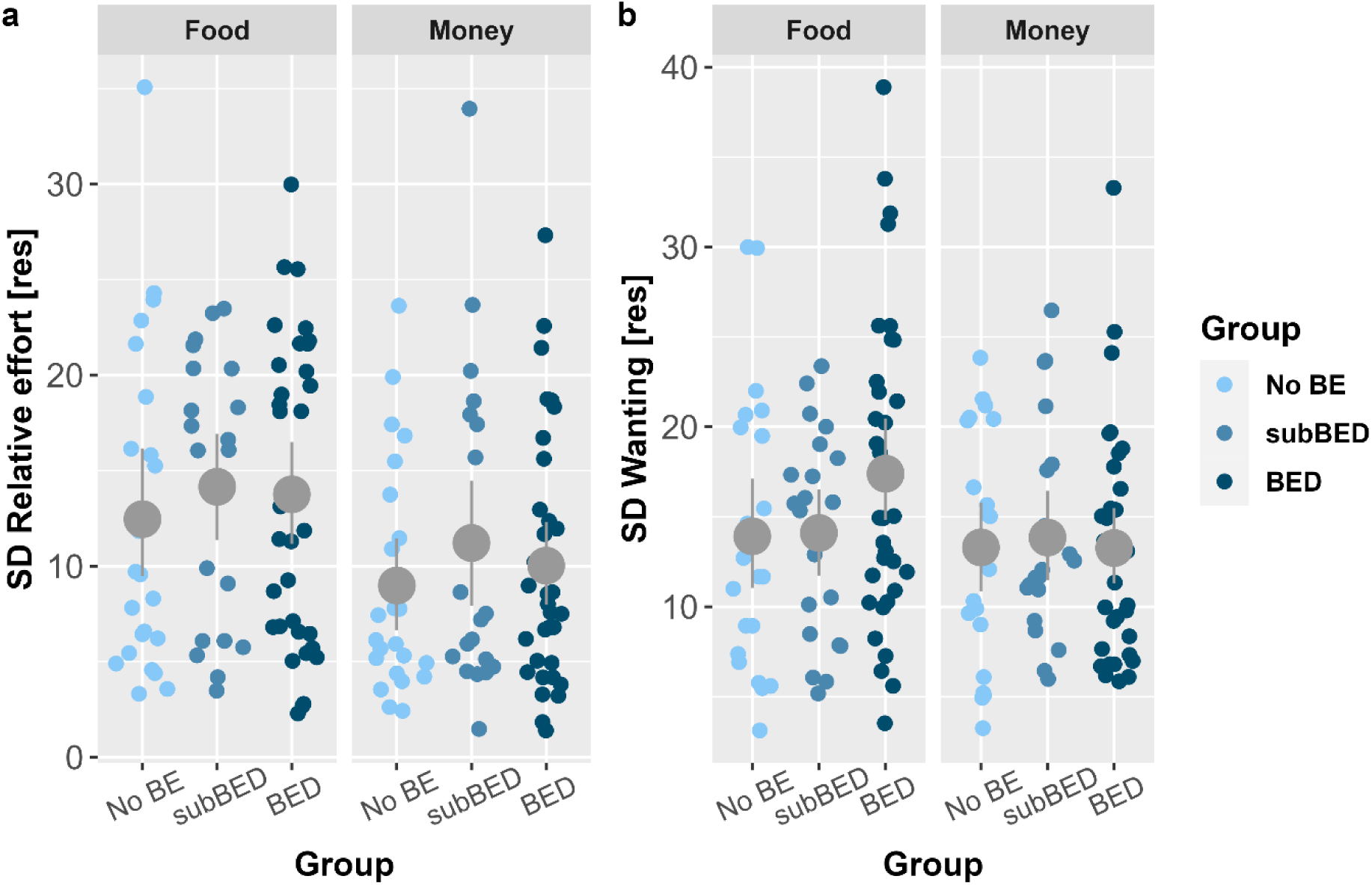
Patients with binge eating disorder (BED) have a higher variability in food wanting ratings during the effort allocation task in the behavioral session. a: No group differences in the standard deviation (SD) of the residuals in trial-wise effort maintenance. b: Patients with BED show higher trial-wise SDs of food wanting ratings (*F*_34,21_=1.48, *p*_boot_ = .024) compared to participants without binge eating (no BE).

### BMI is associated with greater variability of NAcc cue signals

Next, we sought to replicate our previous finding of an increased variability of the NAcc reward response with a higher BMI. In line with previous findings, a higher BMI was associated with more variable NAcc cue responses across trials (*b*=0.11, 95% credible interval, CI [0.03, 0.19], BF_10_=11.1). This association was more pronounced for small rewards (BF_10_=5.3) and money (BF_10_=2.3, Table S3). The main effect of BMI was comparable for certain and uncertain trials (Table S3), but was only dependent on reward magnitude in uncertain trials (BMI×RewM: BF_10_ >20, Table S3).

In addition, we hypothesized that patients with BED would show a higher variability beyond the effects of BMI. Although the variability was numerically higher for subBED (*b*=0.08, 95%CI [−0.11, 0.27]) and BED (*b*=0.10, 95%CI [−0.06, 0.26], BF_10_=0.80), the effects did not reach significance (i.e., 95%CI includes 0). Nevertheless, the posterior probability of a directed (one-sided) test was P(BED>0|data)=88% (BF_10_=7.4, Table S3 for certain and uncertain trials), indicating that the data provide support for an additive effect of BED beyond BMI in the expected direction.

Since patients with BED did not conclusively show enhanced NAcc variability, we sought to replicate the association with disinhibited eating as assessed by the TFEQ (19). To this end, we exchanged the group variable by categories of disinhibited eating. In line with previous findings, variability in NAcc cue responses was higher in participants with high levels of disinhibited eating across the groups (*b*=0.19, 95%CI [0.01, 0.36], BF_10_=4.0, Table S5 for certain and uncertain trials) in addition to the variance explained by BMI as a predictor.

Crucially, including individual SDs of wanting ratings and effort maintenance from the behavioral session showed that higher SDs of wanting were nominally associated with increased variability in NAcc cue responses as indicated by the posterior probability, P(BED>0|data)=89% for the directed test (BF_+0_=8.4; undirected: *b*=0.05, 95%CI [−0.03, 0.14], BF_10_=0.5; driven by uncertain trials, Table S5). In contrast, higher SDs of effort maintenance were not associated with increased variability in NAcc cue responses according to the posterior probability, P(SD_RelEffort>0|data)=12%, (BF+_0_=0.1; undirected BF_10_=0.5, driven by uncertain trials: Table S5).

After replicating the association of higher variability in NAcc cue responses with BMI and disinhibited eating, we next evaluated effects in the dlPFC due to its putative role in top-down control. Again, a higher BMI was associated with more variable dlPFC cue responses across trials (*b*=0.08, 95%CI [0.01, 0.15], BF_10_=2.7). However, patients with BED (*b*=0.04, 95%CI [−0.118, 0.188], BF_10_=0.4) and individuals with higher disinhibited eating (*b*=0.04, 95%CI [−0.13, 0.21], BF_10_=0.5) showed no differences in variability of dlPFC cue responses. In contrast to NAcc cue responses, the association of BMI and variability was only significant in the certain trials (*b*=0.09, 95%CI [0.01, 0.16], BF_10_=2.3, Table S3).

**Figure 3:**
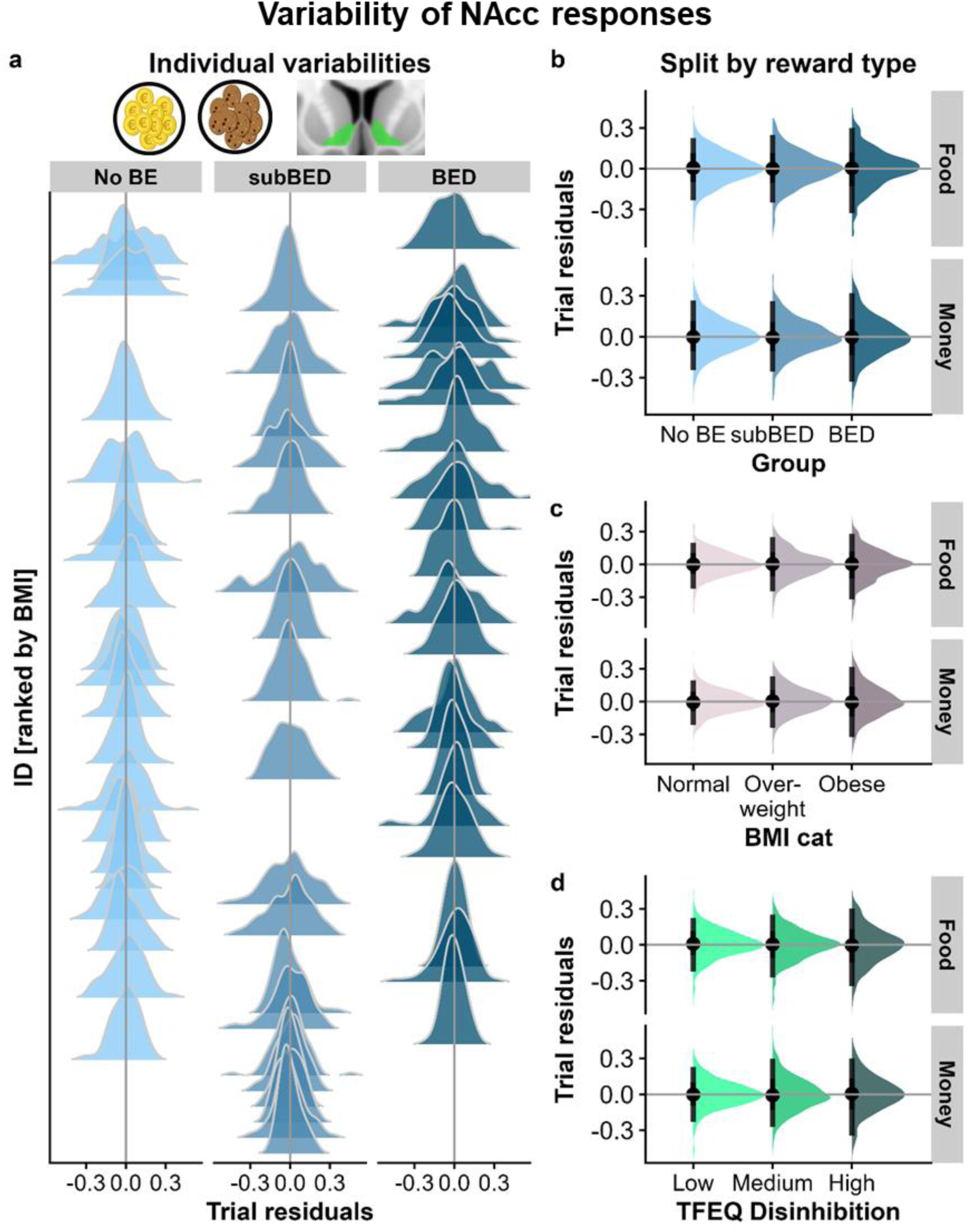
Participants with a higher body mass index (BMI) show elevated variability of anticipatory cue responses in the nucleus accumbens (NAcc). a: Individual distributions of NAcc cue responses ordered by BMI and split by group (no binge eating (no BE), subsyndromal binge eating disorder (subBED), and binge eating disorder (BED). b: Variability of NAcc cue responses is higher in the BED group beyond differences in BMI (directed posterior probability = 88%, *b*=0.10, BF_+0_ = 7.4). However, the undirected 95% CI [−0.06, 0.26] includes 0. Distributions of trial residuals from all participants of the groups. c: Variability of NAcc cue responses is higher in participants with a higher BMI beyond binge eating groups (*b*=0.11, BF_10_=11.1) and more pronounced for money (*b*=0.06 of the interaction, 95%CI [0.01, 0.11], BF_10_=2.3). d: Variability of NAcc cue responses is higher in participants reporting high levels of disinhibited eating beyond differences based on BMI (*b*=0.19, 95%CI [0.01, 0.36], BF10=4.0). Error bars depict 95% percentiles.

### Exploratory and sensitivity analyses

To verify that associations of variability in cue responses with BMI are specific and not explained by mere differences in movement or noise, we performed the same analyses in control regions of the temporal lobe that are largely independent of bottom-up or top-down control signals. There was no association of BMI with cue response variability in the toMTL (b=0.04, 95%CI [−0.04, 0.012], BF_10_=0.3) or pMTL (b=0.02, 95%CI [−0.06, 0.010], BF_10_=0.2) and a weak association in the aMTL (b=0.08, 95%CI [0.00, 0.15], BF_10_=1.3). Notably, all results only changed numerically when including the SD of cue responses in the aMTL to the models, indicating that variability differences in the NAcc and dlPFC are separable from global noise in cue responses (Table S4).

**Figure 4:**
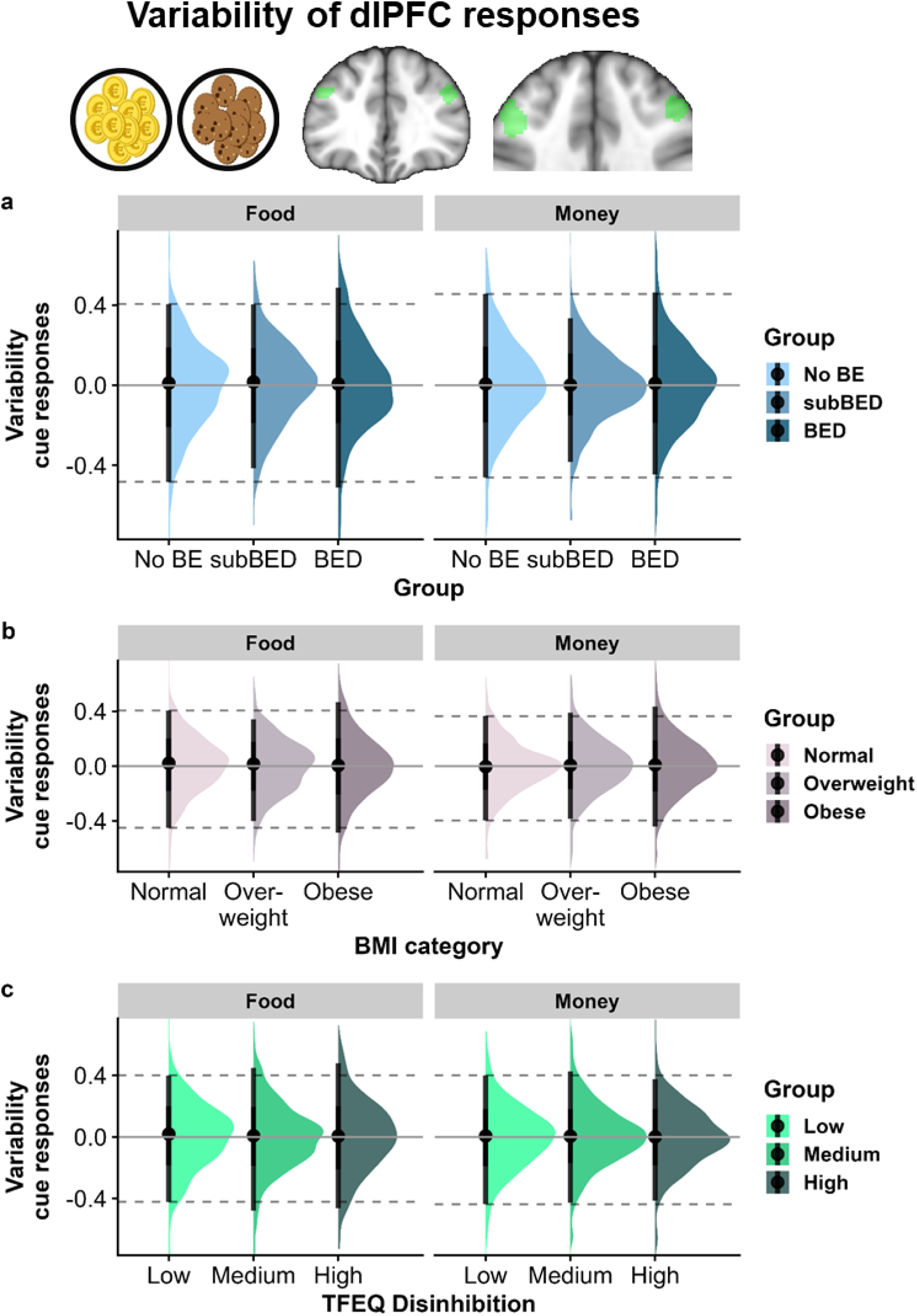
Participants with a higher body mass index (BMI) show elevated variability of anticipatory cue responses in the dorsolateral prefrontal cortex (dlPFC). a: Variability of dlPFC cue responses (distributions of all trial residuals from all participants) does not differ between the binge eating groups. b: Variability of dlPFC cue responses is elevated in participants with higher BMI (*b*=0.08, 95%CI [0.01, 0.15], BF_10_=2.7). c: Variability of dlPFC cue responses does not differ depending on three factor eating questionnaire: disinhibited eating. Error bars depict 95% percentiles.

## Discussion

BED is characterized by repeated episodes of BE with subjective loss of control. Even though such behavior in patients with BED could be seen as an expression of more variable food-related reward behavior compared to individuals without BED (16), most studies have so far focused primarily on potential differences in the amplitude of NAcc responses and its association with symptoms of BE. Here, we showed that variability in food wanting ratings, but not behavioral effort was increased in patients with BED compared to the control groups. We found that increased NAcc variability was positively associated with BMI, as well as TFEQ disinhibition. Against our expectations, this effect was not exacerbated by uncertain effort conditions. In addition to the effect of BMI, we also observed that patients with BED showed slightly elevated variability of NAcc responses as predicted, but the provided level of evidence was inconclusive. To conclude, our findings are the first to highlight the relevance of volatility in subjective experiences of food reward wanting in BED, whilst corroborating the connection between variability in NAcc response, BMI, and food-related disinhibition.

Previous work demonstrated increased variability in NAcc responses is associated with increased BMI and TFEQ disinhibition (19), and that BMI is associated with increased variability in reward learning (38). Here, we replicated this association of variability in NAcc responses with BMI and disinhibition using a different task and focusing on a phase that resembles subjective value signals more narrowly: during anticipation. Whereas Kroemer, Sun (19) measured NAcc variability after milkshake receipt (consummatory response without cues), the current study shows variability in NAcc responses before participants worked for money and food rewards (i.e., by collecting reward points). Since the NAcc is thought to be involved in incentive salience and approach motivation (28, 49), this provides further indication that variability in NAcc responses may be involved in the regulation of reward-directed behavior. Furthermore, BED is often comorbid with obesity (3), and characterized by decreased inhibitory control (50–52). Hence, BMI and disinhibited eating could contribute to the development and maintenance of BED by increasing the volatility of NAcc responses. However, the inconclusive evidence for BED-specific effects in the group-based analysis suggests that these characteristics do not fully account for BE.

Our results extend the literature beyond the more commonly investigated NAcc response amplitude, which has been previously related to weight gain (53, 54) and snacking (55), but not convincingly with current BMI (56–58). Opposed to signal averages, variability in neural and psychological parameters may generally allow for variability in reacting to (food) cues in the environment (59). Our study suggests that heightened variability in NAcc responses may be mechanistically associated with overweight and disinhibited eating. Moreover, since we find a similar association between dlPFC variability and BMI, volatility in top-down processing may additionally contribute to a predisposition for overweight. However, the current study cannot directly disentangle the contribution of top-down processes like inhibitory control. Research directly focusing on response inhibition – for example with the stop-signal or go/no-go task – is thus required to determine whether the association between dlPFC activation and BMI is characterized by greater signal variability beyond lowered amplitudes that have been demonstrated before (15, 60, 61).

The current study could not conclusively support the hypothesis that heightened variability in NAcc responses is a characteristic of BE, not only of an elevated BMI. Instead, we found that patients with BED had a higher variance in their subjective wanting ratings for food compared to subBED or no BE. Although effort to work for reward and wanting are related (62, 63), we previously demonstrated unique correlates of variability in wanting ratings, not effort with BMI in a similar effort task (30)—despite high correlations between wanting rating and behavior on the task (25). Therefore, variability in wanting ratings of food in BED may represent a component of subjective value that is separable from variability in effort. In line with this explanation, variability in wanting and effort were differentially associated with NAcc variability in the current study; whereas variability in wanting ratings corresponded with a higher NAcc variability, more variable effort was associated with *lower* NAcc variability. Since this dissociation was unexpected, future research may help understand how variable wanting ratings of food reward contribute to BED beyond BMI-related variability. One avenue for this could be habitual vs. goal-directed responding (64) as these differ in the degree on which they rely on cost-benefit representations (65).

Despite the notable strengths of our study, the results need to be interpreted under the following considerations. First, our study did not include a measure of variability in food intake, as this would require many repeated sessions to derive good estimates. As such, only limited inferences can be made on the association between variability in NAcc responses and food intake although previous research demonstrated a correlation between variance in *ad libitum* food intake and variability in NAcc responses to milkshake (19). Second, due to the controlled setting under which the experiments took place, we cannot account for variability in NAcc responses that naturally occurs over longer time periods and contexts. Two important fluctuating factors that contribute to BE are metabolic (e.g., 66, 67) and mood states (e.g., 6, 68). Our current study used standardized meals and metabolic states, and no mood induction, deliberately restricting such contributions. Naturalistic designs can better determine whether fluctuations in metabolic and mood states are associated with larger intraindividual variability in reward responses and subsequent BE and we have reported associations of BE with more variable behavior in a gamified reinforcement learning task (38). In this regard, motivational fluctuations should also be taken into account, as they seem to generally influence reward and effort based processing (69). Third, as only women participated in the current study, further research is required to extend the results to men. Furthermore, sex hormones may influence inter- and intra-sex variability in BE (70), which cannot be accounted for in the current study.

To summarize, we have demonstrated that variability in NAcc responses to reward cues is associated with BMI and dietary disinhibition, and that subjective wanting of food cues was more variable in patients with BED compared to participants without BE. Combined, these findings demonstrate that trial-to-trial variability in the reward system as indexed by NAcc responses forms a mechanism that is associated with overweight and uncontrolled eating and that this association goes beyond typically investigated measures of average reward values. Future research should thus focus on how variability in reward processes interacts with other well-established factors contributing to overeating and BE, such as satiation and mood.

## Data Availability

Trial-based data will be shared upon reasonable request to the authors

## Acknowledgement

We thank Dana Wentz, Jacob Schwab, Juliane Zietz, Jennifer Piloth, Lilith Irtel von Brenndorff, Fee Arnold, and Vincent Koepp for help with data acquisition as well as Vanessa Teckentrup for support in preprocessing of the MRI data. The study was supported by the Else Kröner-Fresenius Stiftung grant 2017_A67, and DFG KR 4555/7-1, KR 4555/9-1, and KR 4555/10-1.

## Author contributions

NBK and JS were responsible for the study concept and design. MPN & MH collected data under supervision by NBK. NBK conceived the method and AK & MH processed the data. AK & MH performed the data analysis and NBK contributed to analyses. MH, AK, & NBK wrote the manuscript. All authors contributed to the interpretation of findings, provided critical revision of the manuscript for important intellectual content and approved the final version for publication.

