## supplementalinformation for "Obesity is associated with greater variability of reward signals in the nucleus accumbens"

**Supplemental Information: Body mass index is associated with greater variability of reward signals in the nucleus accumbens**

Mechteld M. van den Hoek Ostende^2,4^, Anne Kühnel^1^, Monja P. Neuser^2^, Jennifer Svaldi^3,4^, & Nils B. Kroemer^1-3, 5*^

^1^	Section of Medical Psychology, Department of Psychiatry and Psychotherapy, Faculty of Medicine, University of Bonn, Bonn, Germany

^2^	Department of Psychiatry and Psychotherapy, Tübingen Center for Mental Health, University of Tübingen, Tübingen, Germany

^3^	German Center for Mental Health (DZPG), partner site Tübingen

^4^	Department of Psychology, Tübingen Center for Mental Health, University of Tübingen, Tübingen, Germany

^5^	German Center for Diabetes Research (DZD), Neuherberg, Germany

**Corresponding author***

Prof. Dr. Nils B. Kroemer,

Venusberg Campus 1, 53127 Bonn, Germany

**Supplemental Methods**

**Grip force effort allocation task**

After a jittered interval, a blue ball appeared at the bottom of the tube to initiate the 24 s effort phase. In certain trials, a red line was drawn across the tube at variable height. This line represented the effort required to collect points so that for each second the participants crossed the line with the ball, they earned reward points. A reward counter at the top right of the screen showed winnings on the current trial. In uncertain trials, however, a muted red area filled part of the tube. During these trials, the difficulty could lay anywhere inside this area and there was no reward counter during these trials. The required effort during the effort phase varied between 64% and 95% (steps of 1% leading to 32 difficulties) of the individual maximum grip force. Each value occurred once for certain, and once for uncertain trials. At the end of the trial, participants received feedback on the number of points they collected on the current trial. After the effort phase of the first session (behavioral), participants additionally indicated on 0-100 visual analog scales (VAS) how much effort they exerted, how much they wanted the reward, and how happy they were at that moment.

The task consisted of 64 randomized trials. During the behavioral session, participants had a single, 30 s break after half the trials, whereas during the neuroimaging session, participants had two 15 s breaks. At the end of the grEAT, participants were paid out their monetary wins in cash, and their food wins in snacks according to their personalized exchange rate. The grEAT was displayed using Psychophysics toolbox v3 (Brainard & Vision, 1997; Kleiner et al., 2007) in MATLAB v2021a (<https://github.com/neuromadlab/Tasks/tree/master/Effort_Allocation_Task>).

**MRI data acquisition and preprocessing**

fMRI data (~35 min, 1500 images) were acquired on a Siemens 3T Prisma scanner with a 64-channel head coil (Details SI). T2*-weighted echo planar images were acquired with a multiband sequence (factor 4) with a short TR of 1.4s and TE of 30 ms (flip angle = 65°, field of view = 220 x 220 mm2) and voxel size of 2 x 2 x 2 mm3. Images covered the whole brain and were acquired with 68 slices with an interleaved slice order. Additionally, an anatomical T1-weighted image was acquired using an MP-RAGE sequence (176 slices, flip angle = 9°, matrix size = 256 x 256, voxel size = 1 x 1 x 1 mm3) as well as a fieldmap to correct for field inhomogeneities (TEshort = 5.19 ms and TElong = 7.65 ms). Data was then preprocessed using fMRIprep (45) and smoothed (6x6x6 m³). For confound correction in first-levels, we extracted the average white matter and CSF signal as well as the six motion regressors.

**MRI firstlevel GLM**

Cue and feedback phases were modeled as events. Work phases were modeled as blocks (22s duration). In addition, we included parametric modulators for magnitude (cue and feedback phase) and difficulty (feedback phase), as well as regressors tracking the exerted force and the change in exerted force for each TR. We included the six movement parameters and average white matter and CSF signal as nuisance covariates.

**BRMS location – scale models**

To determine the effects of group and BMI on the variability in cue responses, we estimated mixed location-scale models (Gaussian link function) with the trial residuals (i.e., deviation from the condition mean) as outcome. In contrast to linear mixed-effects models, location-scale models allow the variance to vary as a function of groups or continuous predictors. Since we used the model only to estimate differences in variability and condition-specific differences in amplitude were already accounted for in the first step, the model for the amplitude only included a random intercept. The model for individual variance terms included the task conditions reward type, reward magnitude, and their interaction. The intercept and main effects of reward type and magnitude were included as random effects. To estimate the effects of BMI (z-standardized) and BE group on trial variability, we included both terms and their interaction with the task conditions as fixed effects. We estimated separate models for the ROIs (NAcc, dlPFC) and each control region (aMTL, pMTL, toMTL). To explore whether effects are driven by certain or uncertain trials, we ran post hoc models on data split by certainty. We used weakly informative normal priors (M=0, SD=0.2) for all regressors modulating the variance and an exponential prior (1) for the SD of the cue variance. Bayesian models were estimated with 4,000 iterations (800 warmup) across 4 chains. Model convergence was evaluated with R ̂ < 1.01. Bayesian models are evaluated using the 95% credible interval that does not include 0 if an effect is significant. In addition, Bayes Factors quantify the evidence for or against the null hypothesis, where BF10 quantifies the evidence for the (undirected) alternative hypothesis and BF01 for the null hypothesis.

**Tables**

Table S1 Bootstrapped F-Tests comparing the variability of wanting and effort maintenance between the no BE and subBED or BED group, respectively.

|  | **subBED (vs. no BE)** | | **BED vs no BE** | |
| --- | --- | --- | --- | --- |
|  | F_(20,21)_ | p_boot_ | F_(34,21)_ | p_boot_ |
| **Uncertain trials: Wanting** | | | | |
| Food | 0.95 |  | 1.48 | **.024** |
| Money | 1.00 |  | 0.97 | .56 |
| Money + Food | 0.98 |  | 1.26 | .11 |
| **Uncertain trials: Effort maintenance** | | | | |
| Food | 1.09 | .39 | 1.13 | .30 |
| Money | 1.51 | .12 | 1.11 | .35 |
| Money + Food | 1.25 | .18 | 1.14 | .28 |
| **Certain trials: Wanting** | | | | |
| Food | 1.01 | .51 | 1.17 | .13 |
| Money | 1.17 | .25 | 1.09 | .45 |
| Money + Food | 1.06 | .39 | 1.13 | .26 |
| **Certain trials: Effort maintenance** | | | | |
| Food | **1.79** | **< .001** | 1.11 | .25 |
| Money | **2.33** | **< .001** | **1.67** | **.007** |
| Money + Food | **1.91** | **< .001** | 1.25 | .076 |

Table S2 Bootstrapped correlations of SDs of wanting and effort maintenance with BMI

|  | r | Lower 95% CI _boot_ | Upper 95% CI _boot_ |
| --- | --- | --- | --- |
| **Uncertain trials** | | | |
| Wanting | .07 | -.08 | .23 |
| Effort | -.06 | -.22 | .08 |
| **Certain trials** | | | |
| Wanting | .06 | -.26 | .02 |
| Effort | -.13 | -.10 | .21 |

Table S3 Results of all models prediction variability of cue responses based on BMI and binge eating

|  | **All trials** | | | |  | **Certain trials** | | | |  | **Uncertain trials** | | | |  |
| --- | --- | --- | --- | --- | --- | --- | --- | --- | --- | --- | --- | --- | --- | --- | --- |
|  | b | 95% CI | BF10 | BF+0 | P>0 | b | CI | BF | BF+0 | P>0 | b | CI | BF | BF+0 | P>0 |
| **Nucleus accumbens** | | | | | | | | | | | | | | | |
| BMI | **0.11** | **0.03; 0.19** | 11.1 | - | - | **0.11** | **0.03; 0.19** | **6.7** |  |  | **0.11** | **0.02; 0.20** | **4.3** |  |  |
| BMI x RewM | **-0.09** | **-0.16; -0.02** | 5.3 | - | - | -0.03 | -0.12; 0.07 | 0.3 |  |  | **-0.17** | **-0.27; -0.08** | **277.5** |  |  |
| BMI x RewT | **0.06** | **0.01; 0.11** | 1.3 | - | - | 0.06 | -0.01; 0.13 | 0.7 |  |  | 0.05 | -0.02; 0.12 | 0.5 |  |  |
| subBED | 0.06 | -0.11; 0.23 | 0.6 | 3.4 | 77% | **0.11** | **-0.06; 0.28** | **0.9** | **9.4** | **90%** | 0.02 | -0.16; 0.20 | 0.5 | 1.5 | 60% |
| subBED x RewM | -0.06 | -0.20; 0.09 | 0.5 |  |  | -0.03 | -0.21; 0.16 | 0.5 | 0.6 | 38% | -0.10 | -0.29; 0.08 | 0.8 | 0.2 | 15% |
| subBED x RewT | 0.04 | -0.08; 0.16 | 0.4 | 3.1 | 75% | 0.03 | -0.13; 0.19 | 0.4 | 1.9 | 65% | 0.05 | -0.11; 0.20 | 0.4 | 2.5 | 72% |
| BED | **0.10** | **-0.06; 0.26** | **0.8** | **7.4** | **88%** | **0.12** | **-0.04; 0.29** | **1.3** | **13.7** | **93%** | 0.07 | -0.11; 0.25 | 0.6 | 3.4 | 77% |
| BED x RewM | -0.04 | -0.17; 0.10 | 0.4 | - | - | 0.02 | -0.15; 0.20 | 0.5 | 1.5 | 60% | -0.07 | -0.26; 0.11 | 0.6 | 0.3 | 22% |
| BED x RewT | 0.04 | -0.07; 0.15 | 0.4 | 2.9 | 75% | 0.06 | -0.09; 0.21 | 0.5 | 3.5 | 78% | 0.03 | -0.12; 0.18 | 0.3 | 1.9 | 66% |
| **dlPFC** | | | | | | | | | | | | | | | |
| BMI | **0.07** | **0.00; 0.15** | **1.4** |  |  | **0.09** | **0.01, 0.16** | **2.3** |  |  | 0.06 | -0.02; 0.14 | 0.6 |  |  |
| BMI x RewM | -0.01 | -0.08; 0.06 | 0.2 |  |  | -0.01 | -0.11; 0.08 | 0.3 |  |  | 0.01 | -0.08; 0.10 | 0.3 |  |  |
| BMI x RewT | 0.01 | -0.04; 0.06 | 0.1 |  |  | -0.01 | -0.09; 0.06 | 0.2 |  |  | 0.04 | -0.04; 0.11 | 0.3 |  |  |
| subBED | -0.03 | -0.19; 0.12 | 0.4 | 0.5 | 34% | 0.02 | -0.15; 0.19 | 0.4 | 1.5 | 59% | -0.09 | -0.26; 0.09 | 0.7 | 0.2 | 16% |
| subBED x RewM | -0.08 | -0.23; 0.06 | 0.7 | 0.2 | 13% | -0.10 | -0.28; 0.08 | 0.9 | 0.2 | 13% | -0.01 | -0.19; 0.17 | 0.5 | 0.8 | 45% |
| subBED x RewT | -0.07 | -0.19; 0.04 | 0.7 | 0.1 | 11% | -0.08 | -0.24; 0.07 | 0.7 | 0.2 | 14% | -0.08 | -0.24; 0.09 | 0.6 | 0.2 | 18% |
| BED | 0.04 | -0.12; 0.19 | 0.7 | 2.2 | 69% | 0.07 | -0.10; 0.23 | 0.6 | 3.8 | 79% | 0.00 | -0.17; 0.17 | 0.4 | 1.0 | 51% |
| BED x RewM | 0.05 | -0.10; 0.19 | 0.7 | 2.9 | 74% | **0.11** | **-0.07; 0.29** | **0.9** | **7.9** | **89%** | -0.04 | -0.22; 0.14 | 0.5 | 0.5 | 33% |
| BED x RewT | -0.02 | -0.13; 0.09 | 0.8 | 0.6 | 36% | -0.08 | -0.23; 0.08 | 0.6 | 0.2 | 16% | 0.04 | -0.12; 0:20 | 0.5 | 2.1 | 68% |
| **aMTL** | | | | | | | | | | | | | | | |
| BMI | 0.08 | 0.00; 0.15 | 1.3 |  |  | 0.07 | -0.01; 0.14 | 0.8 |  |  | 0.07 | -0.02; 0.16 | 0.9 |  |  |
| subBED | -0.05 | -0.21; 0.11 | 0.5 | 0.4 | 27% | -0.05 | -0.22; 0.11 | 0.5 | 0.4 | 26% | -0.07 | -0.25; 0.12 | 0.6 | 0.3 | 23% |
| BED | -0.11 | -0.26; 0.05 | 1.0 | 0.1 | 10% | -0.08 | -0.23; 0.09 | 0.6 | 0.2 | 17% | -0.13 | -0.32; 0.05 | 1.2 | 0.1 | 8% |
| **pMTL** | | | | | | | | | | | | | | | |
| BMI | 0.02 | -0.06; 0.10 | 0.3 |  |  | 0.01 | -0.08; 0.11 | 0.2 |  |  | 0.03 | -0.06; 0.12 | 0.3 |  |  |
| subBED | -0.07 | -0.24; 0.11 | 0.6 | 0.3 | 22% | -0.07 | -0.26; 0.14 | 0.6 | 0.4 | 30% | -0.08 | -0.27; 0.11 | 0.7 | 0.3 | 21% |
| BED | -0.05 | -0.22; 0.11 | 0.5 | 0.3 | 26% | -0.05 | -0.23; 0.13 | 0.5 | 0.5 | 31% | -0.05 | -0.23; 013 | 0.6 | 0.4 | 30% |
| **toMTL** | | | | | | | | | | | | | | | |
| BMI | 0.04 | -0.04; 0.12 | 0.3 |  |  | 0.05 | -0.03; 0.14 | 0.5 |  |  | 0.04 | -0.05; 0.13 | 0.4 |  |  |
| subBED | -0.02 | -0.19; 0.14 | 0.5 | 0.6 | 39% | 0.01 | -0.17; 0.18 | 0.4 | 1.2 | 54% | -0.05 | -0.23; 0.14 | 0.5 | 0.5 | 32% |
| BED | 0.06 | -0.10; 0.23 | 0.6 | 3.3 | 77% | 0.08 | -0.08; 0.25 | 0.7 | 5.2 | 84% | 0.03 | -0.15; 0.22 | 0.5 | 1.8 | 64% |

Table S4 Results when the cue response of the anterior MTL is included in the model as predictor

|  | **All trials** | | | |  | **Certain trials** | | | |  | **Uncertain trials** | | | |  |
| --- | --- | --- | --- | --- | --- | --- | --- | --- | --- | --- | --- | --- | --- | --- | --- |
|  | b | 95% CI | BF10 | BF+0 | P>0 | b | CI | BF | BF+0 | P>0 | b | CI | BF | BF+0 | P>0 |
| **Nucleus accumbens** | | | | | | | | | | | | | | | |
| BMI | **0.11** | **0.03; 0.19** | 8.0 | - | - | **0.11** | **0.03; 0.19** | **7.8** |  |  | **0.11** | **0.03; 0.20** | **5.1** |  |  |
| BMI x RewM | **-0.09** | **-0.16; -0.02** | 5.4 | - | - | -0.03 | -0.12; 0.07 | 0.3 |  |  | **-0.17** | **-0.27; -0.08** | **133.7** |  |  |
| BMI x RewT | **0.06** | **0.01; 0.11** | 1.4 | - | - | 0.06 | -0.01; 0.13 | 0.7 |  |  | 0.05 | -0.02; 0.12 | 0.5 |  |  |
| subBED | 0.06 | -0.10; 0.23 | 0.6 | 3.5 | 78% | **0.11** | **-0.06; 0.27** | **1.0** | **8.7** | **90%** | 0.03 | -0.16; 0.21 | 0.5 | 1.6 | 61% |
| subBED x RewM | -0.06 | -0.20; 0.08 | 0.5 |  |  | -0.03 | -0.21; 0.15 | 0.5 | 0.6 | 38% | -0.10 | -0.28; 0.09 | 0.8 | 0.2 | 15% |
| subBED x RewT | 0.04 | -0.07; 0.16 | 0.4 | 3.3 | 76% | 0.03 | -0.13; 0.19 | 0.4 | 1.8 | 65% | 0.04 | -0.11; 0.20 | 0.5 | 2.5 | 71% |
| BED | **0.10** | **-0.06; 0.26** | **0.9** | **7.7** | **89%** | **0.13** | **-0.04; 0.29** | **1.2** | **13.6** | **93%** | 0.07 | -0.11; 0.25 | 0.6 | 3.7 | 79% |
| BED x RewM | -0.04 | -0.17; 0.10 | 0.4 | - | - | 0.02 | -0.15; 0.20 | 0.5 | 1.5 | 60% | -0.07 | -0.25; 0.11 | 0.6 | 0.3 | 22% |
| BED x RewT | 0.04 | -0.07; 0.15 | 0.4 | 2.9 | 74% | 0.06 | -0.09; 0.21 | 0.6 | 3.5 | 78% | 0.03 | -0.12; 0.18 | 0.4 | 1.9 | 65% |
| **dlPFC** | | | | | | | | | | | | | | | |
| BMI | **0.07** | **0.00; 0.14** | **1.5** |  |  | **0.09** | **0.01, 0.16** | **2.0** |  |  | 0.06 | -0.02; 0.15 | 0.6 |  |  |
| BMI x RewM | -0.01 | -0.08; 0.06 | 0.2 |  |  | -0.01 | -0.10; 0.08 | 0.2 |  |  | 0.01 | -0.08; 0.10 | 0.2 |  |  |
| BMI x RewT | 0.01 | -0.04; 0.06 | 0.1 |  |  | -0.01 | -0.09; 0.06 | 0.2 |  |  | 0.04 | -0.04; 0.11 | 0.3 |  |  |
| subBED | -0.03 | -0.19; 0.12 | 0.4 | 0.6 | 36% | 0.02 | -0.15; 0.19 | 0.4 | 1.4 | 59% | -0.09 | -0.26; 0.08 | 0.7 | 0.2 | 15% |
| subBED x RewM | -0.08 | -0.23; 0.07 | 0.7 | 0.2 | 13% | -0.10 | -0.29; 0.08 | 0.9 | 0.1 | 13% | -0.01 | -0.19; 0.17 | 0.5 | 0.8 | 46% |
| subBED x RewT | -0.07 | -0.19; 0.04 | 0.7 | 0.1 | 10% | -0.08 | -0.24; 0.07 | 0.7 | 0.2 | 15% | -0.07 | -0.24; 0.09 | 0.6 | 0.2 | 19% |
| BED | 0.04 | -0.12; 0.19 | 0.4 | 2.3 | 70% | 0.07 | -0.10; 0.23 | 0.6 | 3.7 | 79% | 0.00 | -0.17; 0.17 | 0.4 | 1.0 | 50% |
| BED x RewM | 0.05 | -0.10; 0.19 | 0.5 | 2.8 | 74% | **0.11** | **-0.07; 0.28** | **0.9** | **7.4** | **88%** | -0.04 | -0.22; 0.14 | 0.5 | 0.5 | 32% |
| BED x RewT | -0.02 | -0.13; 0.09 | 0.3 | 0.6 | 35% | -0.07 | -0.23; 0.08 | 0.6 | 0.2 | 17% | 0.04 | -0.12; 0:20 | 0.4 | 2.1 | 68% |

Table S5 Associations of the variability in Cue responses with disinhibited eating as measured with the Three factor Eating Questionnaire

|  | **All trials** | | | | |  | **Certain trials** | | | |  | **Uncertain trials** | | | |  |
| --- | --- | --- | --- | --- | --- | --- | --- | --- | --- | --- | --- | --- | --- | --- | --- | --- |
|  | b | | 95% CI | BF10 | BF+0 | P>0 | b | CI | BF | BF+0 | P>0 | b | CI | BF | BF+0 | P>0 |
| **Nucleus accumbens** | | | | | | | | | | | | | | | | |
| TFEQ Medium | | 0.03 | -0.13; 0.18 | 0.4 | 1.7 | 63% | 0.08 | -0.08; 0.24 | 0.6 | 4.6 | 82% | -0.01 | -0.18; 0.16 | 0.4 | 0.9 | 47% |
| TFEQ High | | **0.19** | **0.01; 0.36** | **4.0** | **53.7** | **98%** | **0.21** | **0.02; 0.39** | **4.8** | **63.3** | **98%** | **0.17** | **-0.03; 0.36** | **2.0** | **19.7** | **95%** |
| **dlPFC** | | | | | | | | | | | | | | | | |
| TFEQ Medium | | -0.06 | -0.21; 0.09 | 0.5 | 0.3 | 22% |  |  |  |  |  |  |  |  |  |  |
| TFEQ High | | 0.04 | -0.13; 0.21 | 0.5 | 2.0 | 67% |  |  |  |  |  |  |  |  |  |  |

Table S6 Associations of the variability in Cue responses with standard deviations of wanting and effort maintenance during session 1

|  | **All trials** | | | |  | **Certain trials** | | | |  | **Uncertain trials** | | | |  |
| --- | --- | --- | --- | --- | --- | --- | --- | --- | --- | --- | --- | --- | --- | --- | --- |
|  | b | 95% CI | BF10 | BF+0 | P>0 | b | CI | BF | BF+0 | P>0 | b | CI | BF | BF+0 | P>0 |
| **Nucleus accumbens** | | | | | | | | | | | | | | | |
| SD Wanting | **0.05** | **-0.03; 0.14** | **0.5** | **8.4** | **89%** | 0.04 | -0.05; 0.12 | 0.3 | 3.9 | 80% | **0.07** | **-0.02; 0.16** | **1.4** | **13.7** | **93%** |
| SD Effort | **-0.05** | **-0.14; 0.04** | **0.5** | **0.1** | **12%** | -0.04 | -0.13; 0.05 | 0.3 | 0.2 | 20% | **-0.09** | **-0.18; 0.01** | **0.7** | **0.03** | **3%** |
| **dlPFC** | | | | | | | | | | | | | | | |
| SD Wanting | 0.03 | -0.05; 0.11 | 0.3 | 3.8 | 77% | 0.04 | -0.05; 0.12 | 0.3 | 3.8 | 79% | 0.00 | -0.08; 0.08 | 0.2 | 1.0 | 50% |
| SD Effort | -0.01 | -0.09; 0.07 | 0.2 | 0.6 | 39% | -0.02 | -0.11; 0.07 | 0.2 | 0.5 | 32% | 0.02 | -0.06; 0.11 | 0.2 | 2.2 | 69% |
| **aMTL** | | | | | | | | | | | | | | | |
| SD Wanting | 0.02 | -0.07; 0.10 | 0.4 | 1.8 | 65% |  |  |  |  |  |  |  |  |  |  |
| SD Effort | -0.05 | -0.13; 0.04 | 0.2 | 0.2 | 14% |  |  |  |  |  |  |  |  |  |  |
